# Efficacy and safety of vasopressor use in adult patients with traumatic hemorrhagic shock: systematic review

**DOI:** 10.1101/2025.08.10.25333169

**Authors:** Tadaharu Shiozumi, Nobuhiro Sato, Saki Morikawa, Jotaro Tachino, Inaba Mototaka, Naito Hiromichi, Yohei Okada

**Affiliations:** Department of Emergency Medicine, Kyoto Prefectural University of Medicine, Japan; Department of Emergency and Critical Care Medicine, Niigata City General Hospital, Japan; Department of Traumatology and Acute Critical Medicine, Graduate School of Medicine, The University of Osaka, Japan; Department of Emergency, Critical Care, and Disaster Medicine, Okayama University Faculty of Medicine, Graduate School of Medicine, Dentistry, and Pharmaceutical Sciences, Japan; Department of Preventive Services, Kyoto University, Japan

## Abstract

**Background:** Traumatic hemorrhagic shock is a critical condition caused by significant blood loss, leading to impaired tissue perfusion and high mortality. Although vasopressors such as noradrenaline and vasopressin are sometimes administered as adjuncts to fluid and blood product resuscitation, their role remains controversial due to concerns about end-organ perfusion and exacerbation of bleeding.

**Objective:** This systematic review aims to evaluate the efficacy and safety of vasopressor administration in adult patients with traumatic hemorrhagic shock. The review will assess effects on mortality, organ dysfunction, transfusion requirements, adverse events, and tissue perfusion.

**Methods:** We will include randomized controlled trials (RCTs), including cluster-randomized trials, comparing vasopressor use to standard care or no vasopressor in adult trauma patients with hemorrhagic shock. Databases to be searched include CENTRAL, MEDLINE (via PubMed), and Ichushi-Web. The Cochrane RoB 2 tool will be used to assess study quality. Where appropriate, meta-analyses will be conducted using a random-effects model. Subgroup and sensitivity analyses will be performed, and the certainty of evidence will be assessed using the GRADE approach.

**Expected Contribution:** By synthesizing the available evidence, this review will help clarify the role of vasopressors in traumatic hemorrhagic shock, inform clinical practice and guidelines, and identify gaps for future research.

## Background

### Description of the condition

Traumatic hemorrhagic shock is a life-threatening state resulting from severe blood loss, commonly due to blunt or penetrating trauma. It is characterized by inadequate tissue perfusion and cellular oxygenation, leading to organ dysfunction and high mortality. Early recognition and prompt hemostatic resuscitation are essential to restore circulating volume, maintain perfusion, and prevent irreversible shock. Despite advances in trauma care, hemorrhagic shock remains a major contributor to preventable deaths in trauma patients [1].

### Description of the intervention

Vasopressors, such as noradrenaline and vasopressin, are pharmacological agents that increase vascular tone and raise arterial blood pressure. In the context of traumatic hemorrhagic shock, vasopressors are sometimes used as adjuncts to fluid and blood product resuscitation, particularly when hypotension persists despite volume replacement. However, the use of vasopressors in bleeding trauma patients remains controversial, as it may compromise end-organ perfusion or exacerbate ongoing hemorrhage due to vasoconstriction [2].

### How the intervention might work

Vasopressors primarily act through stimulation of α-adrenergic or vasopressin receptors, causing vasoconstriction and increased systemic vascular resistance. Theoretically, this can help maintain perfusion pressure during critical hypovolemia. Noradrenaline also has mild β1-adrenergic activity, which may support cardiac output. Vasopressin, acting independently of adrenergic receptors, may be beneficial in acidotic or catecholamine-resistant states. Nevertheless, excessive vasoconstriction may reduce perfusion to vital organs and increase bleeding by elevating hydrostatic pressure at the site of injury. The net clinical effect of vasopressors in this setting remains uncertain [2, 3].

### Why it is important to do this review

Despite their use in clinical practice, the efficacy and safety of vasopressors in traumatic hemorrhagic shock have not been clearly established [2, 3]. There is no consensus regarding the optimal timing, dosage, or patient selection for vasopressor therapy in this context. Existing studies have reported mixed results, and practice varies widely between institutions and countries [3]. Given the potential for both benefit and harm, a systematic evaluation of the current evidence is needed to guide clinical decision-making, inform practice guidelines, and identify gaps for future research.

### Objectives

To systematically evaluate the efficacy and safety of vasopressor administration in adult patients with traumatic hemorrhagic shock, with a focus on mortality, organ dysfunction, transfusion requirements, adverse events, and tissue perfusion.

## Methods

### Criteria for considering studies for this review

#### Types of studies

We will include all randomized controlled trials (RCTs), including cluster-randomized trials, conducted in actual clinical settings. We will include studies published in English or Japanese. We will exclude simulation or educational studies, as well as unpublished studies and grey literature such as conference abstracts or dissertations.

#### Types of participants

##### Inclusion criteria

We will include adult patients diagnosed with traumatic hemorrhagic shock, with adulthood defined according to the criteria used in each original study. No restriction will be placed on the type of trauma (blunt or penetrating) or the setting of care (e.g., prehospital, in-hospital, or operating room).

##### Exclusion criteria

We will exclude studies that meet the inclusion criteria but fall into one or more of the categories:

- Studies for which full-text articles are not available despite reasonable efforts to obtain them (e.g., inaccessible via institutional subscription or no response from authors).
- Duplicate publications based on the same population or cohort, unless the most complete or recent version can be clearly identified.
- Retracted publications or studies with serious methodological flaws or ethical concerns (e.g., lack of ethics approval, suspected data fabrication).

### Types of interventions

We will consider trials that compared participants who received vasopressors with those who did not. The vasopressor group is defined as patients who received vasopressors such as noradrenaline or vasopressin during the management of traumatic hemorrhagic shock. The control group will be defined according to each original study, such as patients managed without vasopressors, those who received fluid resuscitation alone, or those who received standard care excluding vasopressor use. We will place no restrictions on the type of vasopressor, route of administration, timing of initiation, or dosage. These variables will be considered in subgroup analyses where data permit.

### Types of outcome measures

#### Primary outcomes

1. All-cause mortality at the longest follow-up available.
2. Health-related quality of life (HRQoL), when reported.
3. Short-term mortality, defined as death within 24 hours, 7 days, or 30 days from injury or randomization.

#### Secondary outcomes

4. Length of ICU stay
5. Length of hospital stay
6. Occurrence of organ dysfunction, including multiple organ dysfunction syndrome (MODS) according to definitions used in the included studies.
7. Adverse events related to vasopressor use, such as arrhythmia, myocardial infarction, digital or mesenteric ischemia, acute kidney injury.

### Search methods for identification of studies Electronic searches

We will systematically search the following databases from their inception to the present:

- CENTRAL (Cochrane Central Register of Controlled Trials)
- MEDLINE (via PubMed)
- Ichushi-Web (Japan Medical Abstracts Society)

The search strategies will include a combination of medical subject headings (MeSH terms) and free-text terms related to traumatic hemorrhagic shock, vasopressor agents (e.g., noradrenaline, vasopressin), and randomized controlled trials. The Cochrane Highly Sensitive Search strategy will be adapted to identify randomized controlled trials. We consulted a medical librarian with expertise in systematic reviews to develop and refine the search strategies. We will show our search strategies for CENTRAL, MEDLINE and Ichushi-Web in Appendix 1, Appendix 2 and Appendix 3, respectively. The electronic search strategies were last conducted on 17 June 2025, and will be updated prior to final analysis if necessary.

#### Searching other resources

We will undertake forward and backward citation tracking for key review articles and eligible articles identified through the electronic resources, as appropriate.

#### Data collection and analysis Selection of studies

Two reviewers will independently screen the titles and abstracts of retrieved records. Full texts of potentially eligible articles will be assessed for inclusion. Disagreements will be resolved by discussion or consultation with a third reviewer. The study selection process will be documented in a PRISMA (Preferred Reporting Items for Systematic Reviews and Meta-analyses) flow design [4].

#### Data extraction and management

Two reviewers will independently extract data using a standardized, piloted extraction form. Extracted data will include:

- Study characteristics: author, year, country, design, funding source
- Population: age, sex, inclusion/exclusion criteria, severity of shock
- Intervention: vasopressor type, dose, timing
- Comparator: no vasopressor or standard care
- Outcomes: mortality, organ dysfunction, adverse events

#### Assessment of risk bias in included studies

The Cochrane Risk Bias tool (RoB 2) will be used to assess randomized controlled trials across the following domains:

- Random sequence generation
- Allocation concealment
- Blinding of participants, personnel, and outcome assessors
- Selective outcome reporting
- Other potential sources of bias

Each domain will be rated as “low risk”, “high risk”, or “unclear risk”. Disagreements will be resolved by discussion [5].

#### Measures of treatment effect

- Dichotomous outcome will be summarized using risk ratios (RRs) with 95% confidence intervals (CIs)
- Continuous outcomes will be reported as mean differences (MDs) or standardized mean differences (SMDs) with 95% Cis.
- Time-to-event outcomes will be analyzed using hazard ratios (HRs), when available.

#### Unit of analysis issues

For studies with multiple intervention arms, we will combine relevant arms or use appropriate methods to avoid unit-of-analysis errors. Cluster RCTs will be adjusted for clustering using intracluster correlation coefficients (ICCs) if available.

#### Dealing with missing data

When data are missing or incomplete, study authors will be contacted for clarification. If missing data cannot be obtained, we will report the extent and impact of missingness and assess its implications in sensitivity analyses [5].

#### Assessment of heterogeneity

Statistical heterogeneity will be assessed using the I^2^ statistic and chi-square test. An I^2^ >50% will be considered substantial heterogeneity. Clinical and methodological heterogeneity will also be examined qualitatively.

#### Assessment of reporting biases

If ≥10 studies are included in a meta-analysis, publication bias will be evaluated visually using funnel plots and statistically using Egger’s test [5].

#### Data synthesis

Where appropriate, we will perform meta-analyses using the random-effects model. Fixed-effects models will be used as sensitivity checks. If quantitative synthesis is not feasible. We will conduct a narrative synthesis.

#### Subgroup analysis and investigation of heterogeneity

If data permit, we will perform subgroup analyses by:

- Type of vasopressor
- Timing of administration (early vs. delayed)
- Setting (prehospital vs. in-hospital)

#### Sensitivity analysis

We will conduct sensitivity analyses excluding:

- Studies with high risk of bias
- Studies with imputed or estimated data

### Summary of findings and assessment of the certainty of the evidence

We will use the GRADE approach to assess the overall quality of evidence for each outcome and will present the results in a Summary of Findings table, including key outcomes such as mortality, organ dysfunction, and serious adverse events.

## Declarations Ethical approval

This study is a systematic review protocol based on previously published data and does not involve any new data collection or human participants. Therefore, ethical approval and patient consent are not required.

## Author approval

All authors have read and approved the final manuscript.

## Data availability

All data used in this review will be obtained from publicly available sources.

## Registration

This protocol has been registered with the UMIN Clinical Trials Registry (R000067182).

## Reporting guidelines

The review will be conducted in accordance with the PRISMA-P and PRISMA 2020 guidelines.

## Appendix 1 CENTRAL (Cochrane Central Register of Controlled Trials)

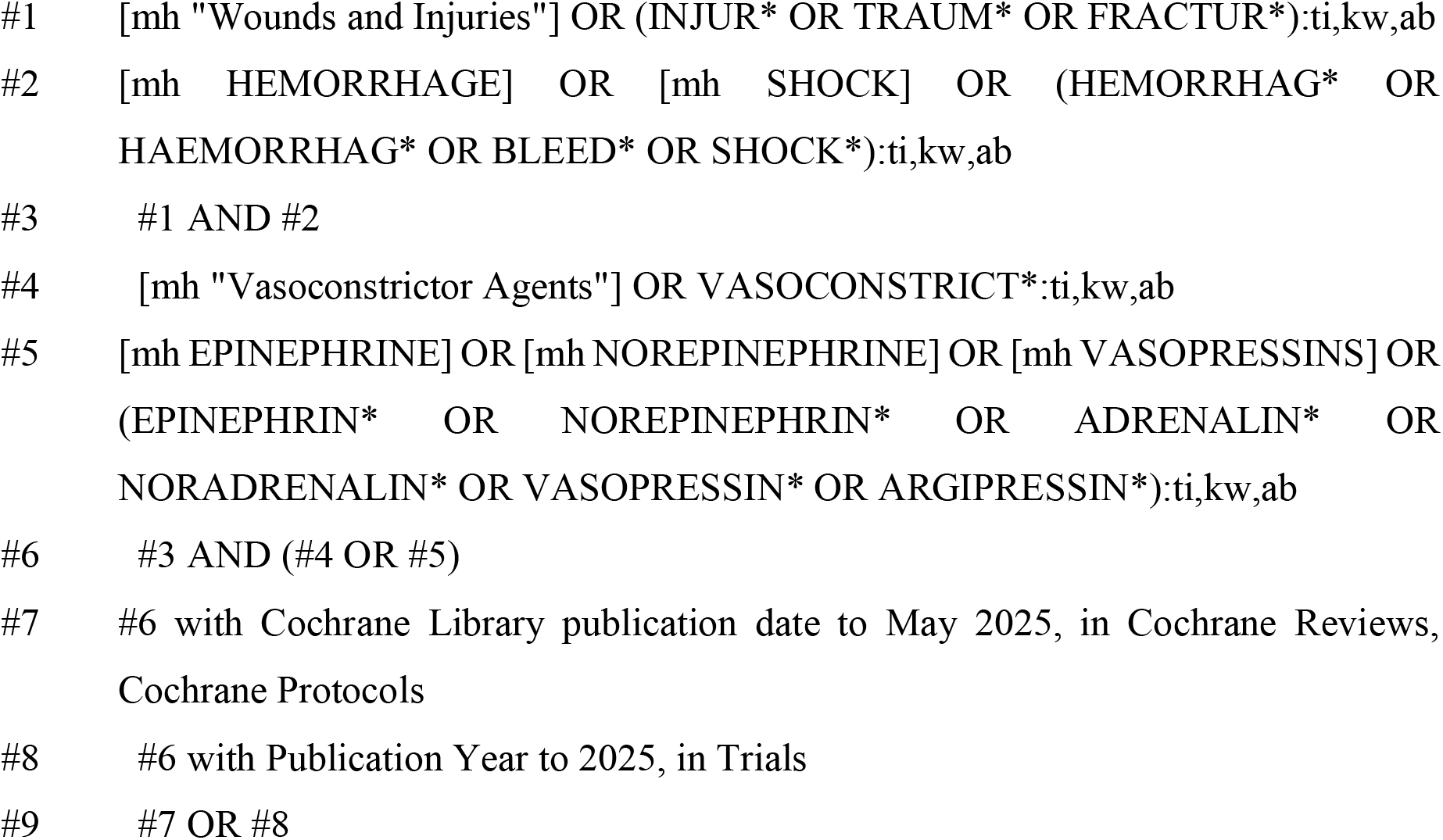

## Appendix 2 MEDLINE (via PubMed)

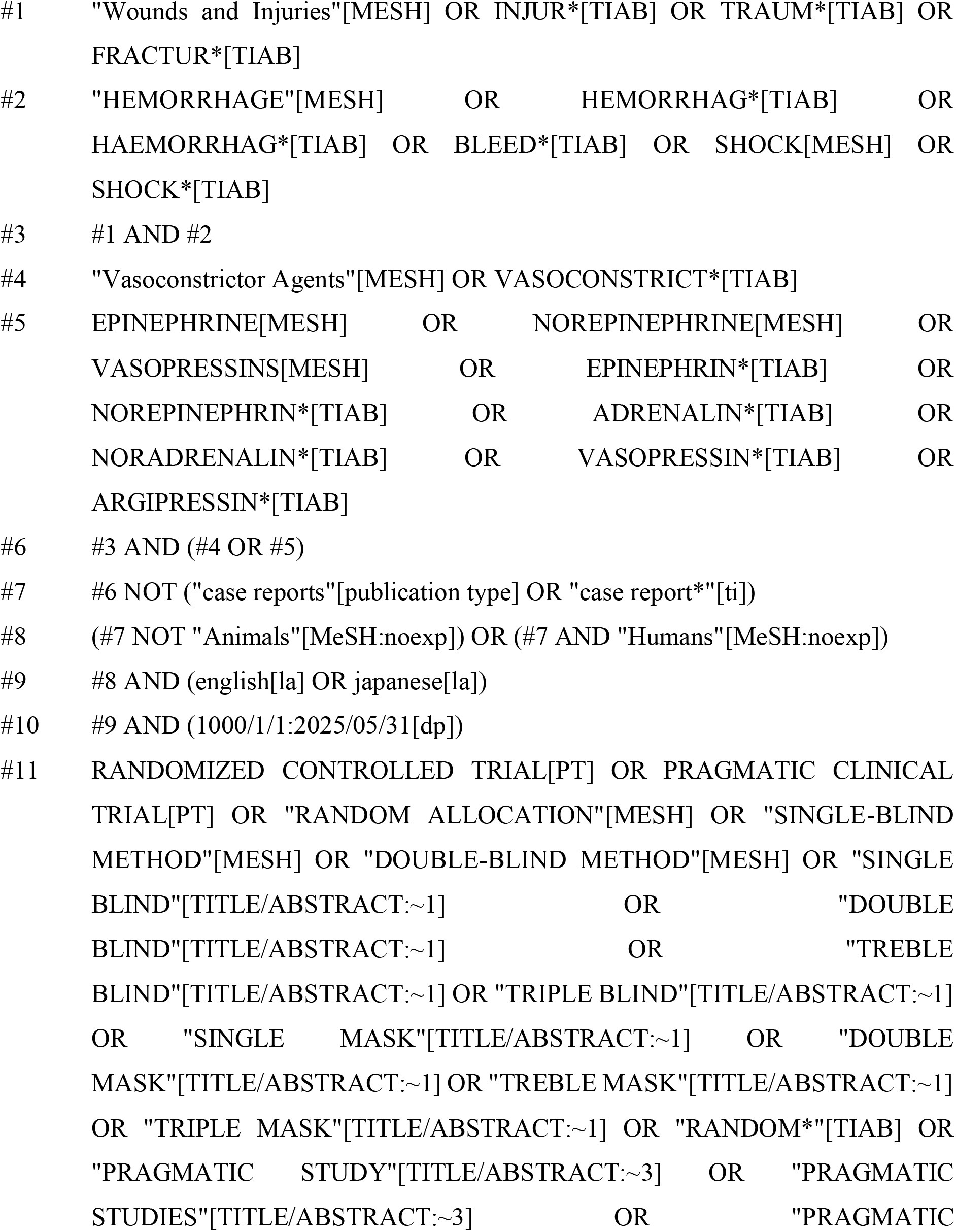
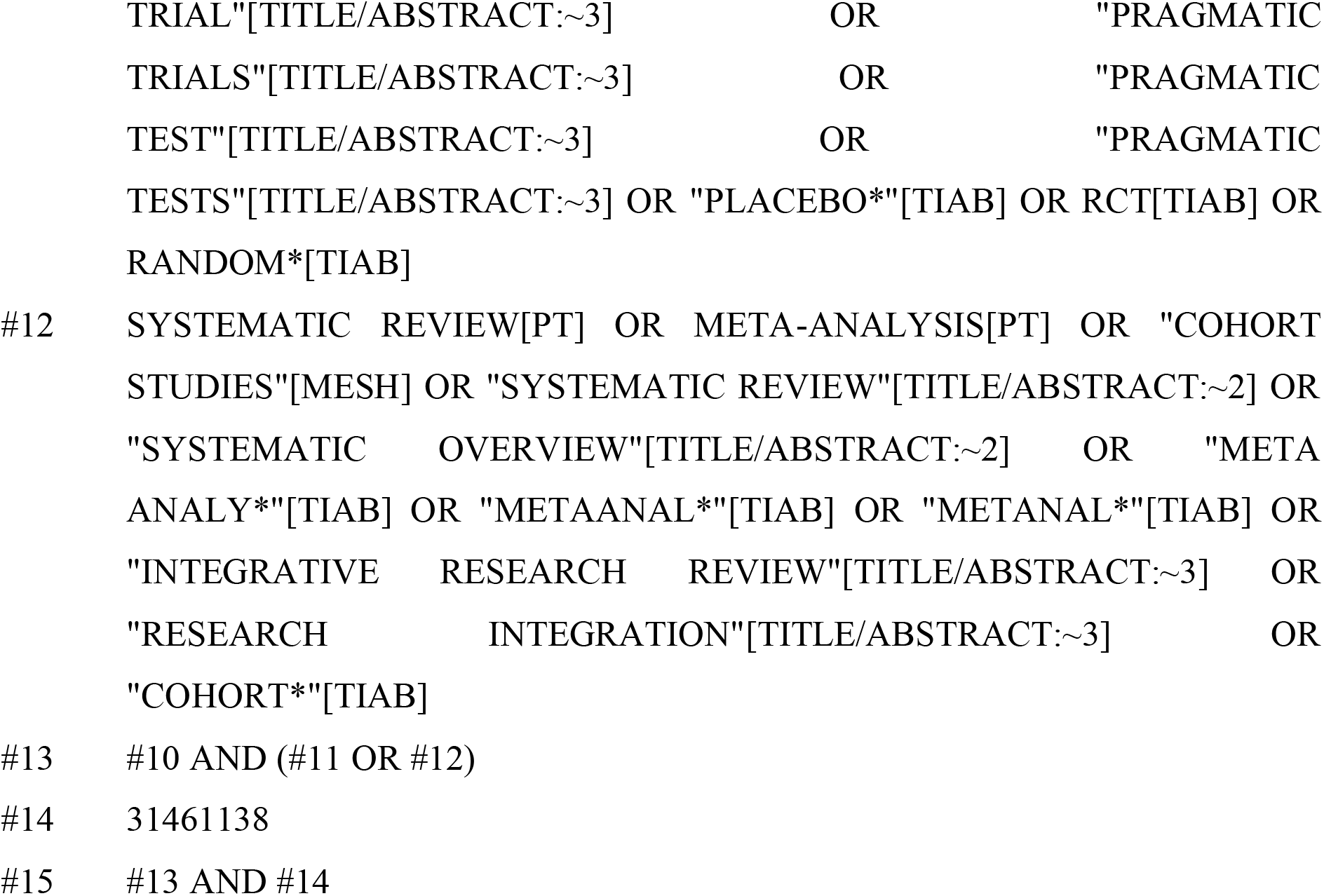

## Appendix 3 Ichushi-Web (Japan Medical Abstracts Society)

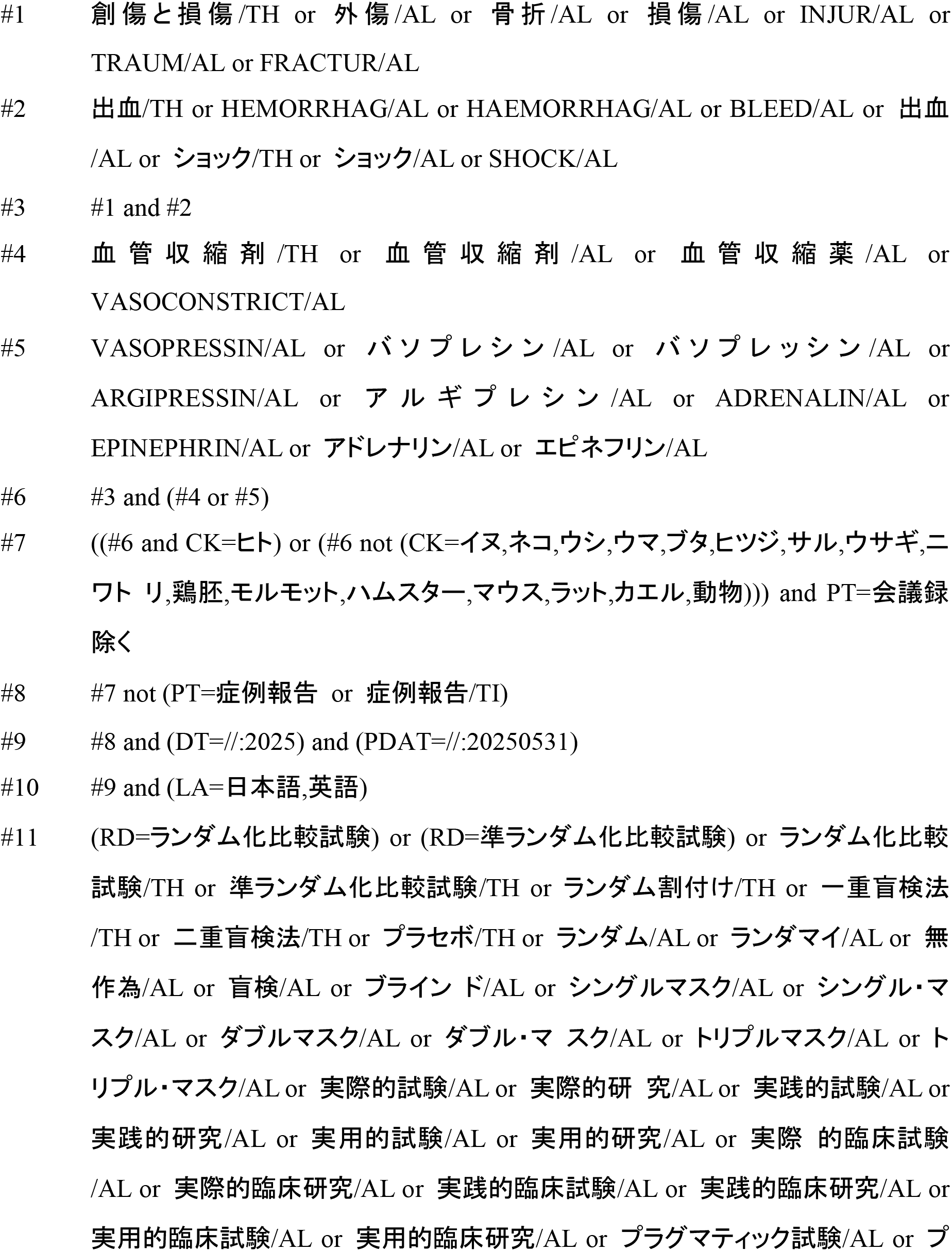
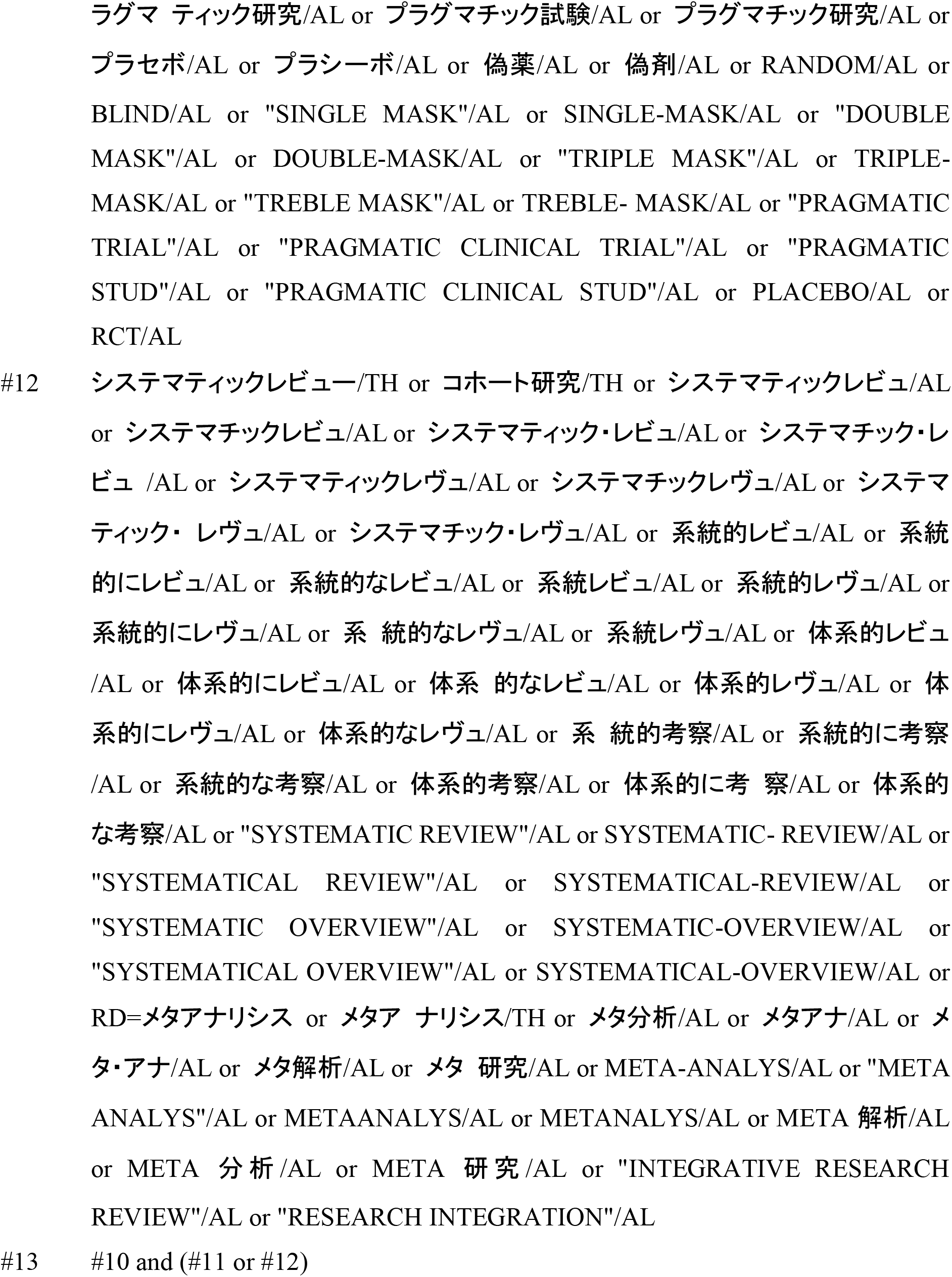

